# Nonresponse adjustment using clinical and perioperative patient characteristics is critical for understanding post-discharge opioid consumption

**DOI:** 10.1101/2021.07.02.21259901

**Authors:** Chris J. Kennedy, Jayson S. Marwaha, P. Nina Scalise, Kortney A. Robinson, Brandon Booth, Aaron Fleishman, Larry A. Nathanson, Gabriel A. Brat

## Abstract

**Background:** Post-discharge opioid consumption is an important source of data in guiding appropriate opioid prescribing guidelines, but its collection is tedious and requires significant resources. Furthermore, the reliability of post-discharge opioid consumption surveys is unclear. Our group developed an automated short messaging service (SMS)-to-web survey for collecting this data from patients. In this study, we assessed its effectiveness in estimating opioid consumption by performing causal adjustment and comparison to a phone-based survey as reference.

**Methods:** Patients who underwent surgical procedures at our institution from 2019-2020 were sent an SMS message with a link to a secure web survey to quantify opioids consumed after discharge. Several patient factors extracted from the EHR were tested for association with survey response. Following targeted learning (TL) nonresponse adjustment using these EHR-based factors, opioid consumption survey results were compared to a prior telephone-based survey at our institution as a reference.

**Results:** 6,553 patients were included. Opioid consumption was measured in 2,883 (44%), including 1,342 (20.5%) through survey response. Characteristics associated with inability to measure opioid consumption included age, length of stay, race, tobacco use, and missing preoperative assessment. Among the top 10 procedures by volume, EHR-based TL nonresponse bias adjustment corrected the median opioid consumption reported by an average of 57%, and corrected the 75th percentile of reported consumption by an average of 11%. This brought median estimates for 6/10 procedures closer to telephone survey-based consumption estimates, and 75th percentile estimates for 3/10 procedures closer to telephone survey-based consumption estimates.

**Conclusion:** We found that applying electronic health record (EHR)-based machine learning nonresponse bias adjustment is essential for debiased opioid consumption estimates from patient surveys. After adjustment, post-discharge surveys can generate reliable opioid consumption estimates. Clinical factors from the EHR combined with TL adjustment appropriately capture differences between responders and nonresponders and should be used prior to generalizing or applying opioid consumption estimates to patient care.

## Introduction

Overprescription of opioids after surgery poses a significant risk to patients and society.^1^ In an effort to standardize prescribing practices and limit overprescribing, many institutions have created guidelines based on expert clinical consensus to help surgeons determine how many opioids to prescribe after a given procedure.^2,3^ While an important first step, these consensus-based guidelines would still result in significant excess opioid prescribing.^2^ Recommendations regarding appropriate prescription sizes benefit from real-world evidence in the form of post-discharge opioid consumption data from patients. The few studies using real consumption data to guide post-surgical prescribing have demonstrated significant clinical improvement in opioid prescribing patterns.^2,4–7^ However, only a few groups have collected this data at scale because it requires significant resources and is labor-intensive to collect.

At our institution, we previously used a phone-based approach to collect post-discharge opioid consumption data from surgical patients. This involved research staff members calling patients following discharge to inquire about opioid consumption.^8^ This survey had an excellent response rate and the accuracy of such telephone-based post-surgical opioid consumption surveys has been validated elsewhere.^8,9^ The consumption data it produced proved useful in guiding many institutional prescribing protocols and in evaluating the validity of existing guidelines and prescribing practices.^2,8,10^ However, the telephone-based method of data collection was not easy to scale to include additional procedures or institutions without a concomitant increase in staffing. In an effort to reduce the manual component of data collection, we developed a new, automated method of data collection. We used a short messaging service (SMS)-to-web system to solicit opioid consumption data from patients. Patients were sent a text message containing a link to a secure web survey on opioid consumption that they could complete on their phone.

Prior studies have shown that several perioperative patient- and procedure-specific factors are associated with opioid consumption after surgery.^11,12^ Some prior studies suggest that these factors are associated with survey response as well.^13–17^ Factors associated with both response and consumption (“confounders”) will distort survey results, rendering estimates of typical opioid consumption non-generalizable. Few prior opioid consumption surveys describe the characteristics of survey respondents in terms of these perioperative characteristics, or adjust for characteristics that may significantly alter survey results.^11,12,18^ In this study we take a comprehensive approach to adjusting for nonresponse bias by describing the characteristics of non-responders using perioperative factors from the electronic health record (EHR) and then adjusting for these characteristics to capture the complex relationship between health system data, survey response, and opioid consumption.

The purpose of this study is to evaluate the validity of our SMS-to-web-based approach for collecting post-discharge opioid consumption data from surgical patients. We used targeted learning, a machine learning-based causal inference methodology, to adjust our web survey data and compared the adjusted consumption data to telephone consumption data that served as a reference standard. In doing so, we found that adjusting for nonresponse--a step that prior studies have generally overlooked--is critical in producing reliable opioid consumption estimates and understanding how consumption should guide evidence-based prescribing guidelines. Our overall framework for understanding surgical opioid use is shown in Figure 1.

**Figure 1.**
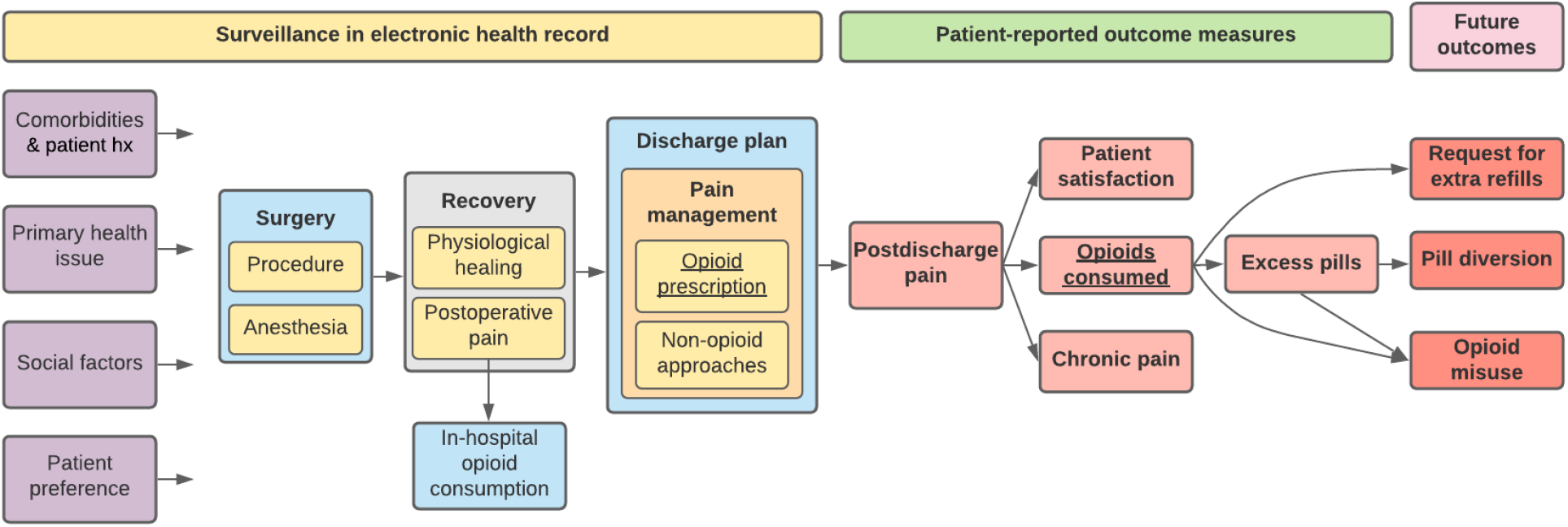
Framework for surgical opioid usage.

## Methods

Following institutional review board (IRB) approval, we created a prospective, single-institution SMS-to-web-based opioid use survey to measure post-discharge opioid consumption among surgical patients.

### Survey-Based and EHR-Based Data Collection

Patients who underwent a surgical procedure at Beth Israel Deaconess Medical Center (BIDMC) from March 1, 2019 to October 31, 2020 were prospectively surveyed after discharge to quantify opioids consumed and to measure satisfaction with pain control. Patients were included if they were 18 years old or greater, spoke English, had a valid cell phone number, and underwent any surgery at BIDMC during the study period. Patients were excluded if they were: trauma patients, patients undergoing multiple surgeries during their index hospitalization, hospitalized for greater than 2 weeks after their surgery, discharged to a rehab facility, had already participated or were readmitted prior to completing the survey, incarcerated patients, or expired patients. An initial SMS message was sent 14 days after discharge, and up to 2 reminder messages at 7-day intervals. Specific survey content is included in Supplemental Table 1. The survey was hosted using Research Electronic Data Capture (REDcap) software.^19^ Patients’ post-discharge opioid consumption was measured in one of two ways: 1) they reported how many pills were remaining from their prescription in the survey, or 2) no opioid prescription was recorded in the EHR.

The primary outcome was the number of opioid pills consumed by the patient after discharge from the surgical procedure, converted to morphine milligram equivalents (MMEs). Initial opioid prescription size, patient demographics, comorbidities, hospitalization status, and surgical- and anesthesia-related characteristics were programmatically extracted from the EHR. The purpose of extracting these EHR-based variables was to comprehensively perform nonresponse bias adjustment of our survey results so that we could more accurately estimate MME consumption. A summary listing of the 40 predictors is provided in Supplemental Table 2.

To provide a reference for comparison for our SMS-to-web survey results, we used a prospective database of opioid consumption data from a prior telephone-based survey at our institution. For this survey, patients who underwent surgery at our institution from October 2017 to June 2018 were called within a month after their surgery and asked how many pills remained. Patients who were still taking opioids were called at 2-week intervals for up to 3 calls in order to record the final tally of pills consumed. Of 3,302 contacted patients, 1,980 (60%) fully answered phone surveys about their post-discharge opioid consumption. Additional details are provided in a previous study.^8^

### Statistical Analysis

Variation in response by EHR characteristics was evaluated using chi-squared tests. Continuous variables were grouped into discrete bins based on common clinical thresholds (e.g. BMI) or based on sample quantiles (e.g. age) so that different subgroups of these variables could be displayed; the Kolmogorov-Smirnov (K-S) test results on raw continuous variables were comparable. Missing predictor data was imputed using generalized low-rank models^20,21^, and missingness indicators were included as predictors after eliminating perfectly collinear indicators.^22^ Additional details on missing data imputation are included in the supplemental information.

Nonresponse bias is a primary threat to survey-based data collection, where estimates of the outcome distribution (opioid consumption in our case) are confounded by variables that are related both to probability of responding to the survey and to the outcome.^23,24^ We conducted a targeted learning (TL)-based double-robust analysis that reduces confounding bias in three ways: 1) inverse probability weights based on a propensity score estimate, 2) an outcome regression adjustment that estimates the relationship between patient characteristics and opioid consumption, and 3) a targeting step that improves estimation using the equation of the specific statistical parameter being studied (i.e. quantiles in our case).^25,26^ We used this TL approach because quantile estimation tasks like the ones we performed in this study (i.e., providing an estimation of the median and 75th percentile of opioid consumption) based on targeted learning has shown efficiency gains equivalent to an increased sample size of 100% or more.^27^ In other words, TL adjustment may improve the accuracy of quantile estimations as much as doubling our sample size would.

Although adjustment can be performed with simpler logistic regression or linear probability models, their linear formulas oversimplify the complex relationship between predictor variables and our two outcomes of interest, opioid consumption and survey response.^28,29^ Accepting this misspecification bias may distort consumption estimates further from the truth, leading to potential patient harm. Machine learning-based estimators have the potential to more accurately capture complex nonlinear relationships between predictor variables and survey response, thereby minimizing residual bias.^30^ We used an ensemble machine learning approach, called the Super Learner algorithm, to evaluate multiple prediction algorithms for their accuracy in capturing these relationships, and to create a weighted average designed to achieve the best accuracy.^31–33^ These modeled relationships were then used to generate quantile estimations using TL as described above. We evaluated the relative importance of predictors by tallying how frequently they were used in decision tree splits of the best performing machine learning algorithm, Bayesian additive regression trees (BART).^34^ Additional estimation details are provided in the supplemental information.

## Results

6,553 surgical patients were surveyed regarding their opioid consumption after discharge. Of these patients, 1,342 (20.5%) responded to the survey and with additional EHR data we were able to measure post-discharge opioid consumption for 2,868 (44%). A data flow diagram of all patients is shown in Figure 2. Baseline characteristics of all included patients are shown in Table 1.

**Figure 2.**
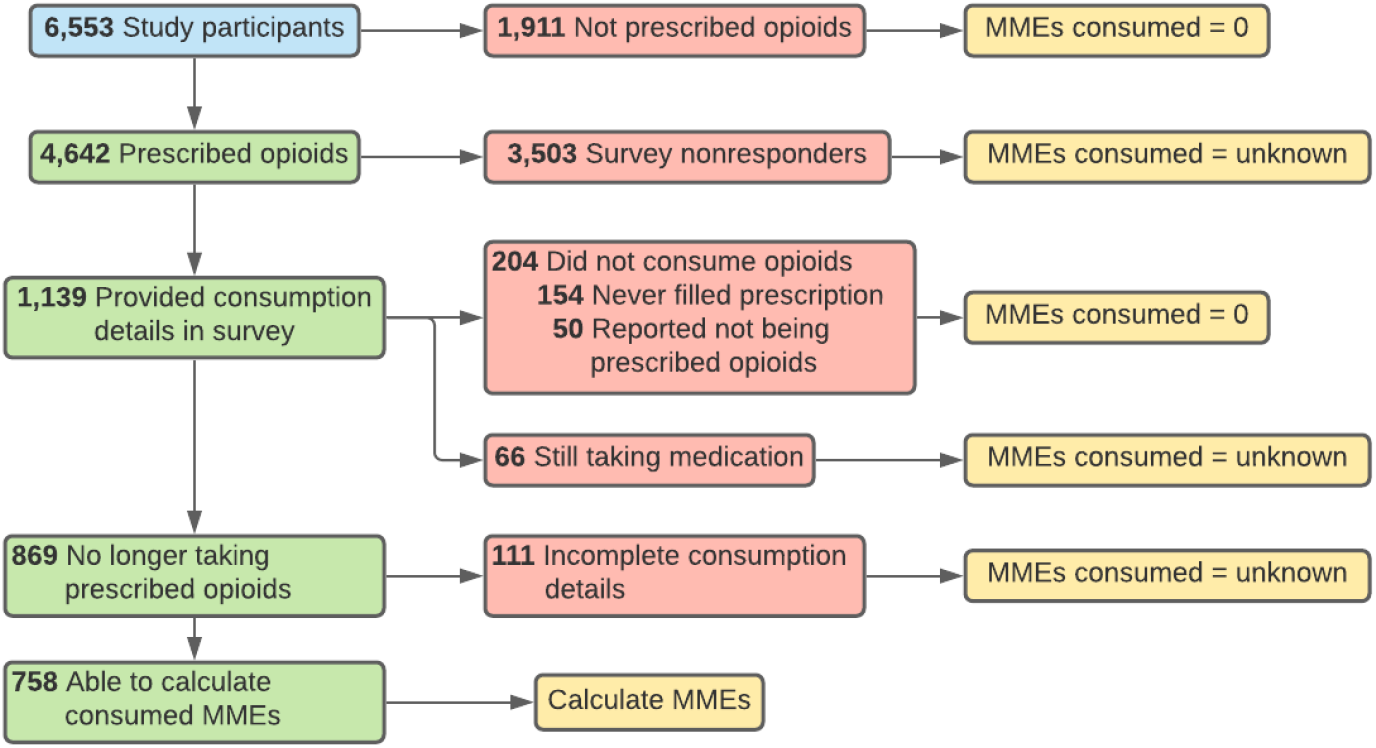
Opioid consumption calculation for study participants.

**Table 1.** Baseline characteristics of study participants: all study participants, survey responders, and those for whom opioid consumption could be measured either through survey response or EHR data.

| Characteristic | Value | Full study<br>(n=6,553) | Survey responders<br>(n=1,342) | Opioid consumption<br>measured<br>(n=2,868) |
| --- | --- | --- | --- | --- |
| Age | 18 - 45 | 24.0% | 19.0% | 21.9% |
|  | 46 - 60 | 31.9% | 33.8% | 30.9% |
|  | 61 - 70 | 26.1% | 31.4% | 27.0% |
|  | 71 - 95 | 18.0% | 15.8% | 20.3% |
| Missing preoperative assessment | No | 86.5% | 89.8% | 87.7% |
|  | Yes | 13.5% | 10.2% | 12.3% |
| Race | White | 72.9% | 80.6% | 74.0% |
|  | Asian | 2.6% | 2.4% | 2.8% |
|  | Black | 11.1% | 6.7% | 9.9% |
|  | Hispanic | 1.9% | 1.1% | 2.0% |
|  | Other | 11.5% | 9.2% | 11.3% |
| Length of stay | Outpatient | 41.7% | 39.9% | 44.6% |
|  | 1 - 2 days | 31.1% | 35.0% | 30.9% |
|  | 3 - 15 days | 27.1% | 25.1% | 24.5% |
| Tobacco use | Never used | 54.9% | 55.9% | 55.2% |
|  | Within past month | 9.2% | 5.9% | 7.4% |
|  | Within 1-12 months | 3.0% | 2.8% | 3.1% |
|  | History | 32.9% | 35.4% | 34.3% |
| Surgical service | Other | 57.4% | 62.4% | 58.4% |
|  | Orthopedic | 30.3% | 28.9% | 28.4% |
|  | Colorectal | 4.8% | 3.3% | 3.6% |
|  | Transplant | 4.3% | 3.4% | 4.7% |
|  | Vascular | 3.2% | 2.0% | 4.9% |
| Opioid drugs prescribed | 0 | 29.0% | 28.2% | 66.3% |
|  | 1 | 68.8% | 70.1% | 32.9% |
|  | 2 or 3 | 2.2% | 1.6% | 0.8% |
| Opioid MMEs prescribed | 0 | 29.1% | 28.3% | 66.3% |
|  | 1 - 99 | 30.6% | 31.5% | 14.8% |
|  | 100 - 299 | 24.1% | 25.1% | 11.7% |
|  | 300+ | 16.3% | 15.2% | 7.2% |

The characteristics of survey responders and nonresponders and a bivariate analysis of select EHR-based clinical factors’ association with survey response are shown in Table 2. Characteristics significantly associated with survey response included age 61-70 years (24.7% response rate, p<0.001), white race (22.6%, p<0.001), length of stay 1-2 days (23.0%, p=0.002) presence of a preoperative assessment visit (21.2%, p<0.001), never smoker status (21.6%, p<0.001), daily alcohol use (27.6%, p<0.001), and American Society of Anesthesiologists (ASA) class 2 (22.5%, p=0.001). Characteristics significantly associated with the ability to measure a patient’s opioid consumption (either through survey pill count or through EHR data indicating no opioid prescription) included age 71-95 years (49.2% consumption measurement rate, p<0.001), outpatient surgery (46.8%, p<0.001), presence of a preoperative assessment visit (44.3%, p=0.018), a history of tobacco use (46.2%, p<0.001), and 0 MMEs consumed in the hospital in the 24 hours prior to discharge (i.e., 0 discharge day MMEs) (52.4%, p<0.001). Using BART analysis, the relative predictive importance of each clinical characteristic is shown in Figure 3a, revealing that age and smoking status are the two most important predictors of response.

**Figure 3.**
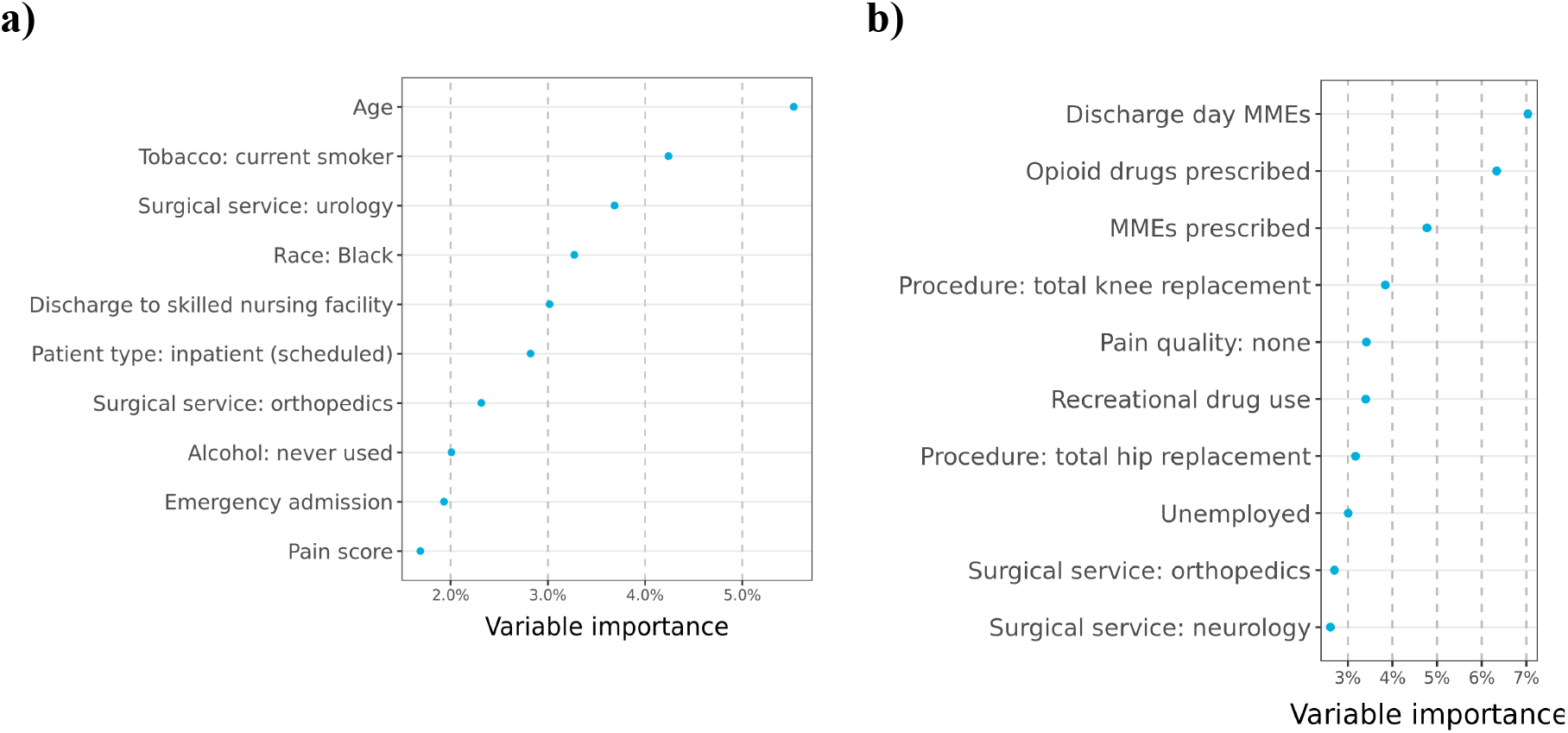
(a) Ranking of clinical and perioperative characteristics most associated with the ability to measure opioid consumption via survey or EHR based on BART, among patients prescribed opioids. (b) Ranking of EHR characteristics most associated with MMEs consumed among patients prescribed opioids. These include random forest-based feature screening to select the top 25 important variables, prior to indicator-encoding categorical variables.

**Table 2.**
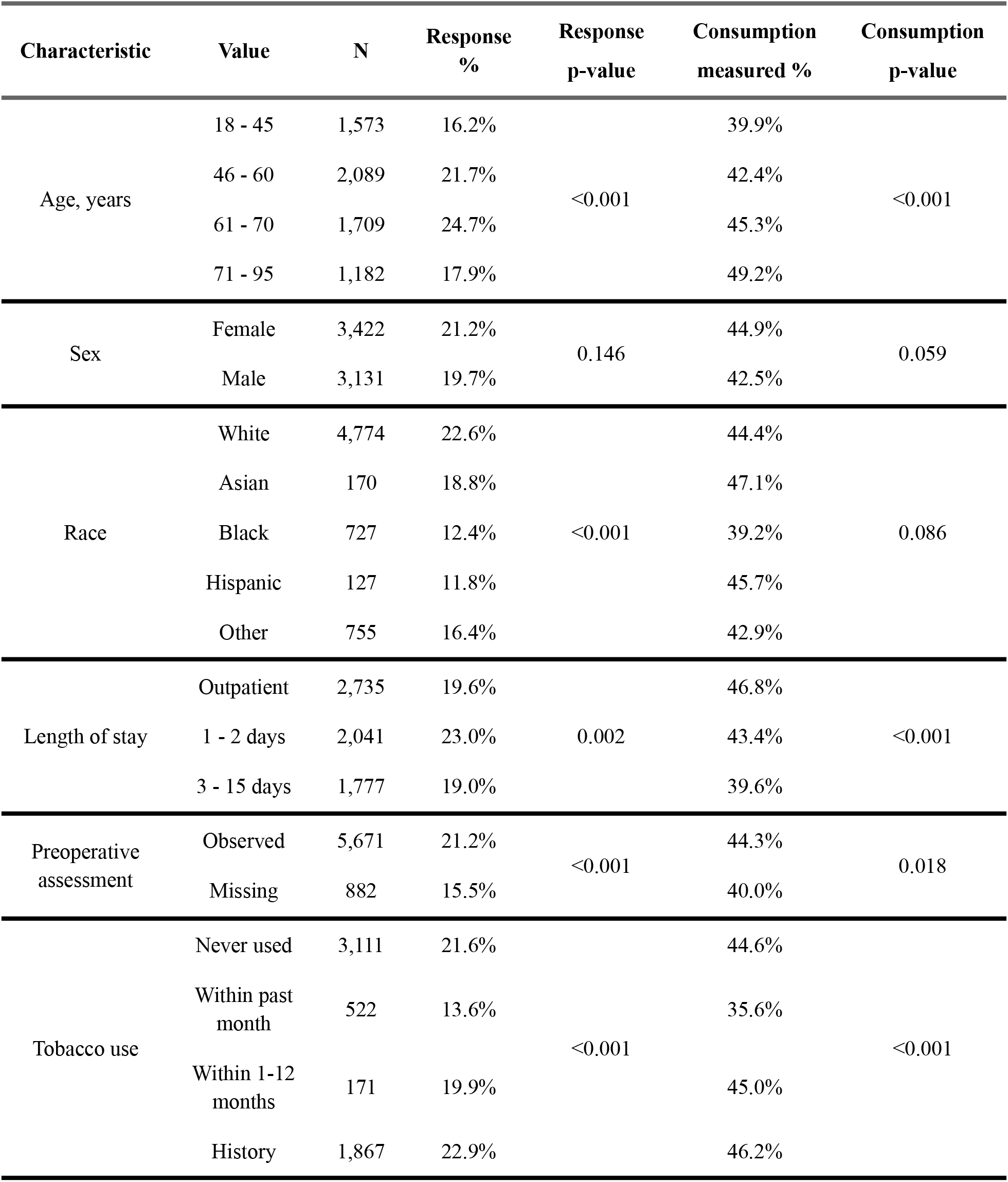
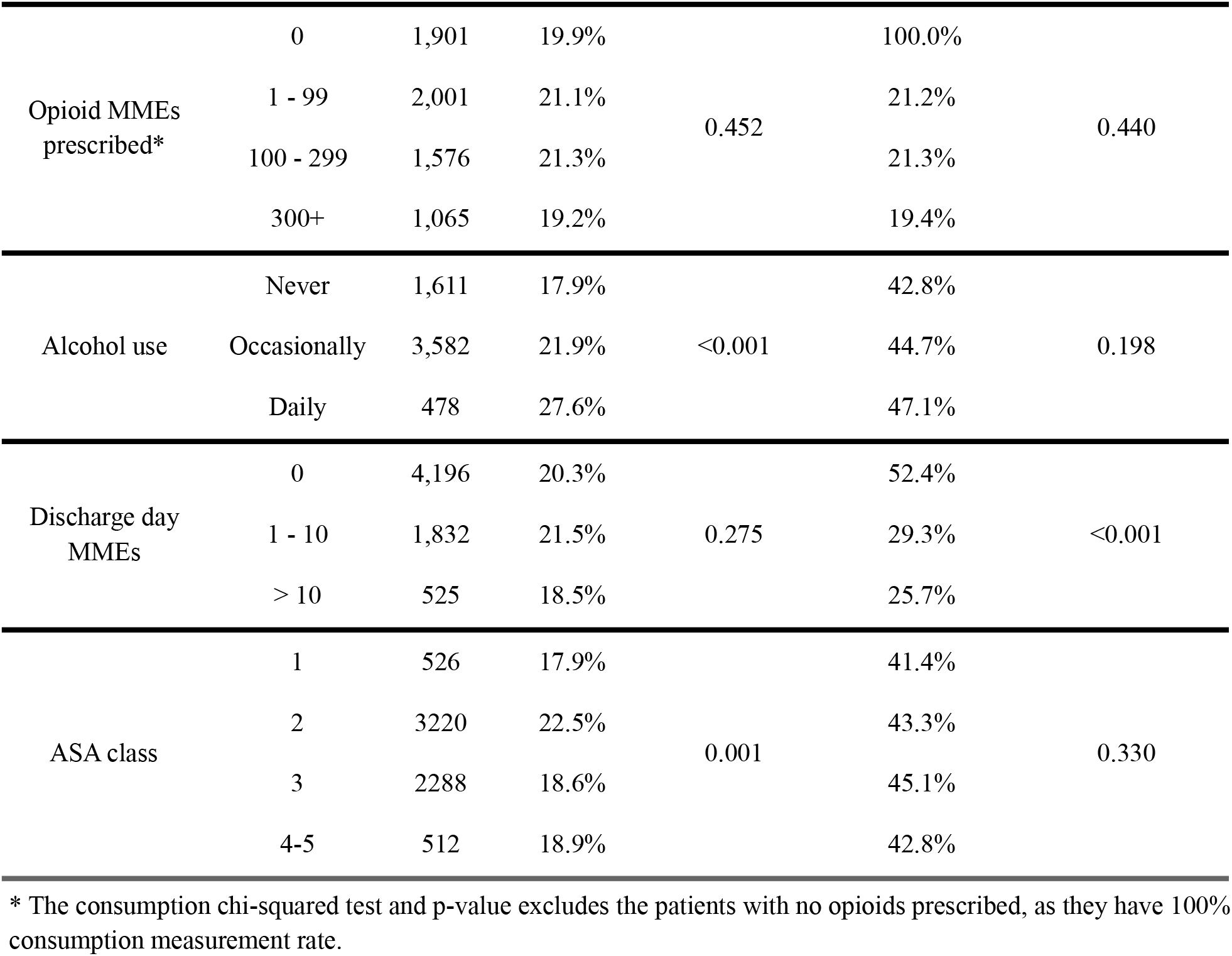
Bivariate analysis of clinical and perioperative factors associated with ability to measure opioid consumption, either through survey response or EHR data.

Some of these predictors of response overlap with predictors of MME consumption, as identified by BART analysis (Figure 3b). In addition, multiple prior studies - including analysis of our telephone survey data - have also shown that age, race, smoking status, and comorbidity burden are strongly associated with MME consumption.^11,12,35^ The association of these clinical and perioperative predictors with both response and consumption suggests potential observed confounding and highlights the importance of EHR-based nonresponse bias adjustment: failing to adjust (or adjusting with only demographic factors) may generate highly biased estimates of opioid consumption, which, if used to inform prescribing guidelines, may lead to patient harm.

Following TL-based nonresponse bias adjustment of our SMS-to-web data, we estimated the median and 75th percentile MME consumption for the 10 most common surgical procedures in our dataset, and compared them against telephone-based estimates (Figure 4). TL nonresponse bias adjustment corrected median MME consumption estimates by an average of 57%, and corrected 75th percentile consumption estimates by an average of 11%, compared to the prior validated telephone survey results. This adjustment brought the median estimates for 6/10 procedures (Figure 4a) closer to telephone survey-based consumption estimates, and brought the 75th percentile estimates (Figure 4b) for 3/10 procedures closer to telephone survey-based consumption estimates. A comparison of demographics of SMS-to-web survey responders and telephone survey responders is shown in Supplemental Table 3.

**Figure 4.**
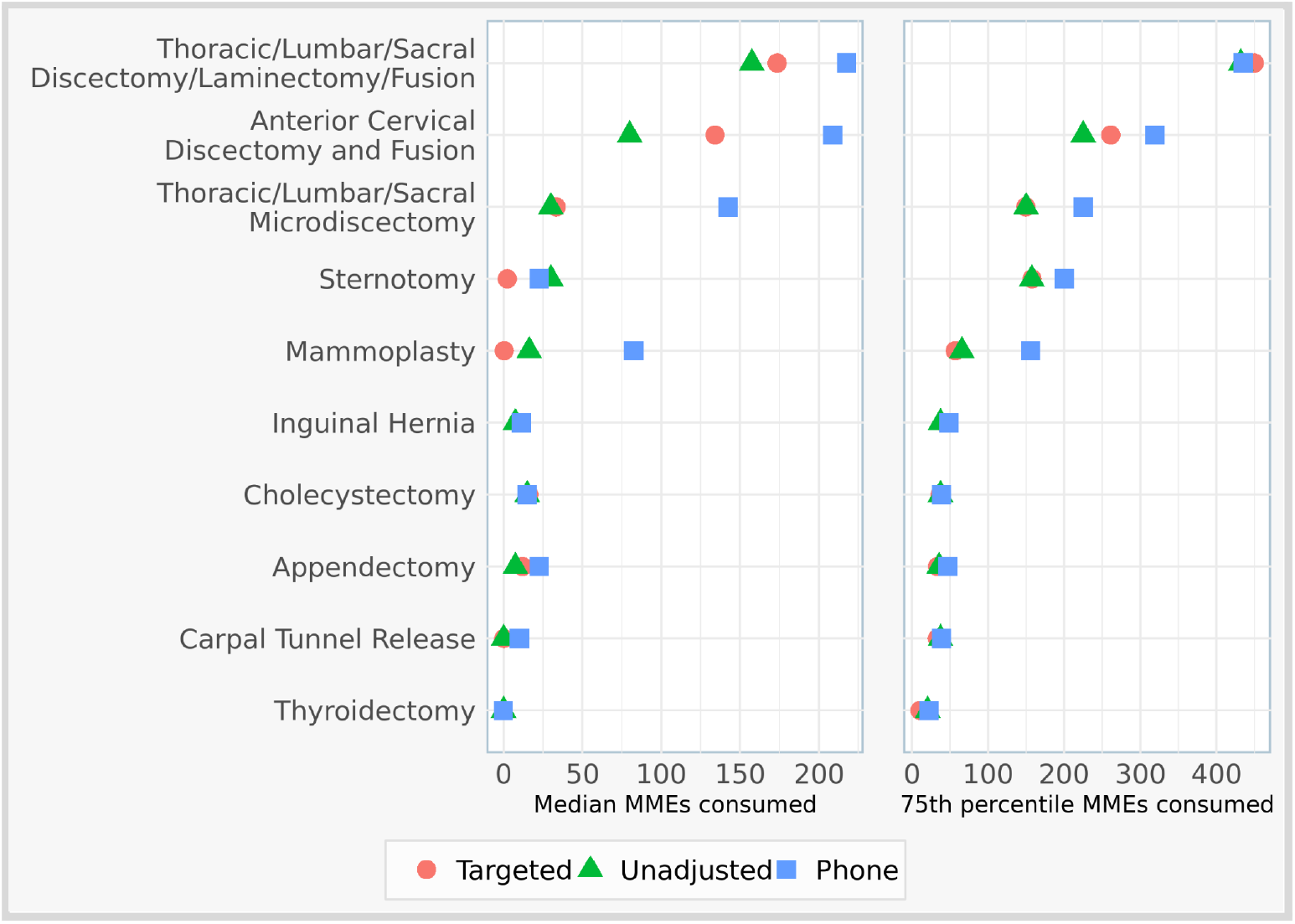
Opioid consumption in MMEs for the 10 most common surgical procedure groups by patient volume, examining the difference between nonresponse bias adjusted and unadjusted survey results, and comparing to the earlier phone survey.

Figure 3 also shows that, across the top procedures, median consumption was only 36% of the median prescription size; 64% of prescribed opioids were not consumed. 75th percentile consumption represented 72% of 75th percentile prescription size. Point estimates of opioid consumption are provided in Supplemental Table 4.

## Discussion

In this study we examined the characteristics of an SMS-to-web survey to measure opioid consumption after discharge amongst post-surgical patients. We found that several clinical and perioperative factors were associated with both survey response and opioid consumption, suggesting that unadjusted survey measurements were biased. Using clinical factors found in the EHR to adjust for nonresponse bias significantly changed typical opioid consumption estimates from the survey data. The difference in opioid consumption estimates using unadjusted versus adjusted survey data was clinically significant, which indicates that EHR-based nonresponse bias adjustment is a critical step in producing reliable opioid consumption that can be used to inform prescribing guidelines.

### Prior Opioid Consumption Surveys

Nonresponse bias is a challenge for any survey-based data collection.^23^ Clinical studies often overlook the issue of nonresponse and do not adjust for it;^7,36^ failing to do this prior to interpreting survey results can lead to biased results and non-generalizable conclusions. Few opioid consumption studies have examined any factors associated with nonresponse. A recent study measuring post-discharge opioid consumption among patients undergoing orthopedic and urologic surgery using an SMS-based survey found that the presence of comorbidities were associated with nonresponse, however nonresponse adjustment was not performed.^18^ To our knowledge, no opioid consumption surveys have adjusted their consumption estimates for nonresponse bias.^11,12,18^ The accuracy and generalizability is largely unknown for surveys in which nonresponse bias has not been examined.

### Importance of EHR-based Adjustment

We found that several EHR-based clinical and perioperative factors were associated with both opioid consumption and survey response in our study, suggesting that unadjusted estimates of opioid consumption may be biased. Multiple prior studies confirm this overlap: age, race, smoking status, and comorbidity burden are associated with MME consumption.^11,12,35^ The association of these clinical and perioperative predictors with both response and consumption is confounding and may lead to inaccurate opioid consumption estimates if not incorporated into adjustment, and may adversely affect patient care if used to guide opioid prescribing recommendations. Nonresponse bias adjustment using these factors significantly revised the median and 75th percentile opioid consumption estimates for most procedures. Several prior clinical surveys on other topics have uncovered associations between EHR-based clinical factors and survey response as well.^13–17^We also used EHR data to improve the completeness of our consumption data, instead of relying on survey-reported opioid consumption alone. Patients who did not respond to the survey but were not prescribed any opioids in the EHR were counted as consuming zero MMEs in our final dataset. In merging survey data with EHR-based patient outcome data we were able to increase the number of patients for whom opioid consumption could be measured.

### Importance of Machine Learning-Based Adjustment

Traditional parametric methods for nonresponse bias adjustment such as logistic regression are limited in their ability to correct for this bias because they do not fully capture the complex patterns that differentiate responders and nonresponders.^28,29^ The bias that is accepted by these simple models may distort opioid consumption estimates so that they are further from the truth, leading to harm by misinforming prescribing guidelines. Ensemble machine learning, as we used in this study, can reduce estimation bias compared to logistic regression by relying on algorithms that best predict the outcome and response variables.^30,37^ In observational studies at risk of confounding bias, it is a best practice to use machine learning-based adjustment to generate the least biased estimates. Future surgical and opioid research should incorporate these recent methodological developments.^38^

### Limitations

While our prior telephone-based survey results served as the standard of comparison for our SMS-to-web survey results, it is important to note that there is no “gold standard” survey method for measuring opioid consumption; telephone-based consumption data is vulnerable to nonresponse bias as well. Furthermore, there may have been background trends that influenced opioid prescribing habits and opioid consumption at our institution between 2017-2018 when the telephone data was collected, and 2019-2020 when the SMS-to-web data was collected. As our study and prior work has identified, one of the strongest predictors of opioid consumption is prescription size.^11^ The difference between SMS-to-web consumption estimates and telephone consumption estimates, even after nonresponse adjustment, may be partially explained by these two phenomena. Nonetheless, we chose to use our telephone-based consumption data as a reference for comparison since it is a commonly used method for collecting opioid consumption data in prior studies, had a high response rate of 60% in our study, and its accuracy has been validated by previous work demonstrating that it aligns closely with in-person based consumption estimates.^8,9,11,12^ While our SMS-based survey had a comparatively lower response rate as expected,^18,39,40^ the automated nature of this survey method may limit recall bias. We were able to automatically contact patients on a weekly basis after discharge, while studies using telephone and other labor-intensive survey methods have waited up to 12 months after discharge to survey patients on their pain and opioid consumption after surgery.^7^

## Conclusion

In conclusion, survey nonresponse bias adjustment using EHR-based clinical factors is a critical step in understanding post-discharge opioid requirements for surgical patients. Since many clinical factors associated with nonresponse overlap with factors related to opioid consumption, adjustment using these factors is essential. The ease of data collection using a nonresponse adjusted SMS-to-web survey may enable large-scale opioid consumption data to guide future opioid prescribing guidelines. This survey method may be used to collect other patient-reported post-discharge outcomes as well, provided it is adjusted using clinical factors associated with nonresponse or identified as potential confounders through causal diagrams.^41^

## Data Availability

Data unfortunately cannot be shared due to protected health information.

## Acknowledgments

This work was conducted with support from CRICO/Risk Management Foundation of the Harvard Medical Institutions and a Blavatnik Biomedical Accelerator Pilot Grant of Harvard University.

