## Supplementary material for "Nonresponse adjustment using clinical and perioperative patient characteristics is critical for understanding post-discharge opioid consumption"

### Supplemental Study Details

#### Missing data imputation

Missing values were imputed using generalized low-rank models (GLRM), a generalization of principal component analysis that linearly decomposes and reconstructs the original dataset while reducing random variation (noise) in the variable measurements.[1] GLRM hyperparameters were optimized via a grid search based on reconstruction error on a 25% holdout set.[2] The hyperparameter grid consisted of 75 combinations: number of components $\in$ {36, 38, 40}, X regularization $\in$ {none, quadratic, L1}, Y regularization $\in$ {none, quadratic, L1}, X gamma $\in$ {0, 4, 8}, Y $\in$ {0, 4, 8}. The optimal hyperparameters were: components = 40, X regularization = quadratic, Y regularization = quadratic, X gamma = 8, Y gamma = 4. A performance comparison of GLRM-based imputation versus median/mode imputation is provided in Supplemental Table 5.

#### Targeted learning estimation

The SuperLearner library for the outcome regression (prediction of MMEs consumed, among those prescribed opioids) was: the outcome mean, stratification on prescribed MMEs and discharge day MMEs, ordinary least squares (OLS), lasso, and random forest. The latter three algorithms were tested with all predictors and with predictors restricted to those linearly correlated with the outcome at a p-value < 0.1. The SuperLearner library for the propensity score estimation (measurement of opioid consumption) was: the outcome mean, ordinary least squares (OLS), logistic regression, lasso, random forest, and Bayesian additive regression trees (BART). The latter five algorithms were tested with all predictors and with predictors restricted to those linearly correlated with the outcome at a p-value < 0.1.

### References

1. Udell M. Generalized Low Rank Models. Boyd S, Mackey L, Van Roy B, eds. Published

online 2015.
http://search.proquest.com/dissertations-theses/generalized-low-rank-models/docview/2507949861/se-2

2. Kennedy CJ, Mark DG, Huang J, van der Laan MJ, Hubbard AE, Reed ME. Development of an ensemble machine learning prognostic model to predict 60-day risk of major adverse cardiac events in adults with chest pain. *medRxiv*. Published online 2021.

**Supplemental Table 1.** SMS-to-web survey details.

*Web Survey Instrument*

1. On what part of your body was the surgery? (More than 1 may apply. Choose 1.)

    a. Head or Neck (brain, skull, ears, face, mouth, thyroid, parathyroid, throat, carotid)
    b. Back (spine, vertebrae)
    c. Chest (lungs, breast, esophagus, heart)
    d. Abdomen (stomach, intestines, appendix, gallbladder, colon, hernia, liver, pancreas, transplant, kidney, prostate, bladder)
    e. Leg or Arm (toes, foot, ankle, knee, hip, fingers, hand, wrist, elbow, axilla/underarm, shoulder)
    f. Genitals and Buttock (penis, vagina, abscess hemorrhoid, fistula, fissure, pilonidal cyst)
    g. Blood Vessels (angiogram, aorta, vascular repair)

Opioids are pain medications such as oxycodone, Vicodin, Percocet, hydrocodone, Dilaudid, and tramadol. Questions about opioids refer to these types of medications.

2. If you were prescribed an opioid, have you stopped taking your prescription?
    a. I was not prescribed an opioid pain medication
    b. No, I am still taking the opioid medication
    c. I am no longer taking the opioid pain medication (I may still be taking other pain medications)
    d. I never filled the opioid prescription given to me

[If Q2 = c, skip to Q4. If Q2 = a or d, end immediately.]

3. May we text you back in a week to see if your pain has improved?
    a. Yes
    b. No

[Survey completed]

==========

[Questions 4 - 9 are repeated for each opioid-class drug prescribed, between 0 and 3 times.]

4. You were prescribed {number} {medication-name} {medication-strength} dose). Is this correct?
    a. Yes
    b. No

[If Q4 = Yes, skip to Q6]

5. Please tell us what drug, dose (mg), and number of pills you were prescribed.
    a. [Free text input]

6. Did you receive any refills for this prescription?
    a. Yes
    b. No

7. Was this prescription a pill or liquid?
    a. Pill
    b. Liquid

[If Q7 = Liquid, skip to Q9]

8. How many of these pills do you have left?
    a. [Free text input]

[Skip to Q10]

9. Please estimate the percent of liquid you have left.
    a. [Free text input] (%)

=============

10. How would you rate your overall pain control after discharge home from the hospital?
    a. Excellent
    b. Manageable
    c. Intolerable

11. How would you describe your pain?
    a. More than expected
    b. As expected
    c. Less than expected

12. How well did your providers explain how much pain you experienced?
    a. Very well
    b. Reasonably well
    c. Not well at all

13. Have you used other medications for pain including but not limited to: (Please check all that apply)
    a. Tylenol (acetaminophen)
    b. Aspirin
    c. Motrin (ibuprofen)
    d. Aleve (naproxen)
    e. Neurontin (gabapentin)
    f. Lyrica (pregabalin)
    g. Other
    h. None of the above

[If Q13 != Other, Skip to Q15]

14. Please List Other Medication
    a. [Free text input]

15. Prior to surgery, I was taking medications for: (Please check all that apply)
    a. Anxiety
    b. Depression
    c. Sleep
    d. ADHD or ADD
    e. Drug dependence
    f. None of the above

16. For pain prior to surgery, were you taking
    a. Opioids
    b. Non-opioid medications

17. How did you manage your leftover opioid prescription?
    a. Kept them in an unlocked space

b. Kept them in a locked space
c. Took them to dropboxes (police stations and pharmacies) and/or to collection days
d. Flushed them
e. Threw them in the garbage
f. Combined them with coffee grounds, kitty litter or dirt and put in garbage
g. Didn't have excess medication
h. Took them to my doctor
i. Other

*SMS Message Content*

Please help BIDMC fight the opioid crisis. Click on the link to fill out our survey on patient opioid use.

LINK.

Reply to this text with STOP to opt out of future communications.

*SMS Message Timing*

Text messages were sent at 1pm Eastern Time.

**Supplemental Table 2.** Summary listing of predictor variables.

| | Variable | Type | Unique values | Missing rate | Mode | Mean | Median | Min | Max |
|---|---|---|---|---|---|---|---|---|---|
| **Opioids** | | | | | | | | | |
| 1 | discharge_day_mme | Continuous | 105 | 0.0% | 0 | 3 | 0.0 | 0.00 | 65.0 |
| 2 | opioid_drugs_prescribed | Integer | 4 | 0.0% | 1 | 1 | 1.0 | 0.00 | 3.0 |
| 3 | prescribed_mme | Continuous | 166 | 0.2% | 0 | 140 | 75.0 | 0.00 | 7,680.0 |
| **Demographics** | | | | | | | | | |
| 4 | age | Integer | 78 | 0.0% | 64 | 56 | 58.0 | 18.00 | 95.0 |
| 5 | age_group | Ordinal int. | 4 | 0.0% | (45,60] | | | | |
| 6 | employment_status | Categorical | 3 | 13.5% | Occupation | | | | |
| 7 | gender | Categorical | 2 | 0.0% | F | | | | |
| 8 | hispanic_ind | Categorical | 5 | 0.3% | N | | | | |
| 9 | race_group | Categorical | 5 | 0.0% | White | | | | |
| **Comorbidities** | | | | | | | | | |
| 10 | alcohol | Ordinal int. | 3 | 13.5% | Occasionally | | | | |
| 11 | bmi | Continuous | 1,784 | 0.3% | 26.63 | 29 | 28.2 | 8.52 | 67.9 |
| 12 | current_smoker | Binary | 2 | 0.0% | 0 | 0 | 0.0 | | |
| 13 | daily_narcotic_use | Binary | 2 | 13.5% | 0 | 0 | 0.0 | | |
| 14 | hx_opioid_abuse | Binary | 2 | 0.0% | 0 | 0 | 0.0 | | |
| 15 | recreational_drugs | Binary | 2 | 13.5% | 0 | 0 | 0.0 | | |
| 16 | tobacco | Ordinal int. | 4 | 13.5% | Never used | | | | |
| **Surgical** | | | | | | | | | |
| 17 | prim_surg_svc | Categorical | 12 | 10.0% | ORTHO | | | | |
| 18 | wound_class | Categorical | 5 | 0.5% | CL | | | | |
| **Anesthesia** | | | | | | | | | |
| 19 | anesth_type | Categorical | 7 | 0.1% | GEN | | | | |
| 20 | asa_class | Integer | 5 | 0.1% | 2 | 2 | 2.0 | 1.00 | 5.0 |
| 21 | asa_class_e | Binary | 2 | 0.0% | 0 | 0 | 0.0 | | |
| 22 | pain_quality | Categorical | 8 | 13.5% | None | | | | |
| 23 | pain_score | Integer | 11 | 13.5% | 0 | 2 | 0.0 | 0.00 | 10.0 |
| **Hospital** | | | | | | | | | |
| 24 | case_type | Categorical | 3 | 0.1% | S | | | | |
| 25 | ibax_adm_type | Categorical | 3 | 0.3% | EL | | | | |
| 26 | ibax_disch_disp | Categorical | 5 | 0.3% | 01 | | | | |
| 27 | inout_cd | Categorical | 2 | 0.3% | O | | | | |
| 28 | length_of_stay | Integer | 16 | 0.0% | 0 | 2 | 1.0 | 0.00 | 15.0 |
| 29 | los_group | Ordinal int. | 3 | 0.0% | Outpatient | | | | |
| 30 | pat_type_bidmc | Categorical | 3 | 0.1% | INP | | | | |
| 31 | pat_type_sched | Categorical | 4 | 0.1% | OP | | | | |
| 32 | payor_grp | Categorical | 16 | 0.3% | MR | | | | |
| **Missing** | | | | | | | | | |
| 33 | miss_anesth_type | Binary | 2 | 0.0% | 0 | 0 | 0.0 | | |
| 34 | miss_asa_class | Binary | 2 | 0.0% | 0 | 0 | 0.0 | | |
| 35 | miss_bmi | Binary | 2 | 0.0% | 0 | 0 | 0.0 | | |
| 36 | miss_inout_cd | Binary | 2 | 0.0% | 0 | 0 | 0.0 | | |
| 37 | miss_prescribed_mme | Binary | 2 | 0.0% | 0 | 0 | 0.0 | | |
| 38 | miss_prim_surg_svc | Binary | 2 | 0.0% | 0 | 0 | 0.0 | | |
| 39 | miss_tobacco | Binary | 2 | 0.0% | 0 | 0 | 0.0 | | |
| 40 | miss_wound_class | Binary | 2 | 0.0% | 0 | 0 | 0.0 | | |

**Supplemental Table 3.** Demographic comparison between phone and SMS survey responders.

| Characteristic | Value | Phone | SMS | P-value |
|---|---|---|---|---|
| Age, years | 18 - 45 | 22.6% | 17.9% | 0.0001 |
| | 46 - 60 | 29.9% | 33.4% | |
| | 61 - 70 | 28.3% | 31.7% | |
| | 71 - 98 | 19.2% | 16.9% | |
| Sex* | Female | 47.4% | 54.3% | <0.0001 |
| | Male | 50.7% | 45.7% | |
| Race | Asian | 2.2% | 2.5% | 0.0617 |
| | Black | 9.1% | 6.8% | |
| | Other | 10.8% | 10.8% | |
| | White | 77.9% | 79.9% | |
| Body mass index | < 18.5 | 0.9% | 0.8% | 0.5643 |
| | 18.5 - 24.9 | 23.0% | 26.7% | |
| | 25 - 29.9 | 27.9% | 34.8% | |
| | 30+ | 29.9% | 37.7% | |
| | Missing | 18.3% | 0.1% | |

*The sex response option “other” was included in the phone survey instrument but not in the SMS-recruited version of the survey instrument, so that row is not included in this table.

**Supplemental Table 4.** Opioid consumption in MMEs for the top 10 surgical procedure groups by volume, examining the difference between nonresponse bias adjusted and unadjusted survey results, and comparing to the earlier phone survey.

| | | Median | | | | 75th percentile | | | |
|---|---|---|---|---|---|---|---|---|---|
| **Surgical approach** | **n** | **Prescribed** | **Survey Response** | **Targeted** | **Phone** | **Prescribed** | **Survey Response** | **Targeted** | **Phone** |
| Thoracic/Lumbar/Sacral Discectomy/Laminectomy/Fusion | 393 | 300 | 157 | 173 | 217 | 450 | 432 | 449 | 435 |
| Anterior Cervical Discectomy and Fusion | 150 | 225 | 80 | 134 | 208 | 300 | 225 | 261 | 318 |
| Thoracic/Lumbar/Sacral Microdiscectomy | 128 | 225 | 30 | 33 | 142 | 228 | 150 | 149 | 225 |
| Sternotomy | 464 | 160 | 30 | 2 | 22 | 300 | 157 | 157 | 200 |
| Mammoplasty | 173 | 90 | 16 | 0 | 82 | 150 | 65 | 56 | 155 |
| Inguinal Hernia | 289 | 45 | 7 | 9 | 11 | 75 | 37 | 42 | 48 |
| Cholecystectomy | 382 | 37 | 15 | 16 | 15 | 75 | 37 | 37 | 38 |
| Appendectomy | 151 | 37 | 7 | 11 | 22 | 56 | 35 | 33 | 46 |
| Carpal Tunnel Release | 145 | 0 | 0 | 0 | 10 | 37 | 37 | 33 | 38 |
| Thyroidectomy | 122 | 0 | 0 | 0 | 0 | 37 | 20 | 10 | 22 |

**Supplemental Table 5.** Imputation performance comparison of generalized low-rank models with median/mode imputation. The average percent reduction in error across all variables was 46%.

| Variable | Missingness (%) | Error GLRM | Error Std. | Percent reduction |
|---|---|---|---|---|
| employment_status | 13.5 | 0.04 | 0.34 | 89.5 |
| alcohol | 13.5 | 0.37 | 0.37 | 0.0 |
| daily_narcotic_use | 13.5 | 0.77 | 0.25 | -204.0 |
| recreational_drugs | 13.5 | 0.36 | 0.36 | 0.0 |
| tobacco2 | 13.5 | 0.91 | 0.45 | -101.1 |
| pain_quality | 13.5 | 0.09 | 0.34 | 72.2 |
| pain_score | 13.5 | 1.03 | 3.97 | 74.2 |
| prim_surg_svc | 10.0 | 0.15 | 0.70 | 78.0 |
| wound_class | 0.5 | 0.07 | 0.28 | 75.8 |
| hispanic_ind | 0.3 | 0.10 | 0.16 | 38.2 |
| ibax_adm_type | 0.3 | 0.01 | 0.15 | 91.5 |
| ibax_disch_disp | 0.3 | 0.04 | 0.31 | 86.5 |
| inout_cd | 0.3 | 0.73 | 0.69 | -5.5 |
| payor_grp | 0.3 | 0.24 | 0.75 | 68.1 |
| bmi | 0.3 | 1.09 | 6.83 | 84.0 |
| prescribed_mme | 0.2 | 92.9 | 241.3 | 61.5 |
| asa_class | 0.1 | 0.08 | 0.86 | 91.1 |
| anesth_type | 0.1 | 0.04 | 0.26 | 83.5 |
| case_type | 0.1 | 0.02 | 0.09 | 82.8 |
| pat_type_bidmc | 0.1 | 0.00 | 0.52 | 100.0 |
| pat_type_sched | 0.1 | 0.02 | 0.61 | 97.1 |
